# The association between dysnatraemia during hospitalisation and post COVID-19 mental fatigue

**DOI:** 10.1101/2022.07.26.22277990

**Authors:** Gerardo Salvato, Elvira Inglese, Teresa Fazia, Francesco Crottini, Daniele Crotti, Federica Valentini, Giulio Palmas, Alessandra Bollani, Stefania Basilico, Martina Gandola, Giorgio Gelosa, Davide Gentilini, Luisa Bernardinelli, Andrea Stracciari, Francesco Scaglione, Elio Clemente Agostoni, Gabriella Bottini

**Affiliations:** Department of Brain and Behavioral Sciences, University of Pavia, Pavia, Italy; Cognitive Neuropsychology Centre, ASST “Grande Ospedale Metropolitano” Niguarda, Milano, Italy; NeuroMI, Milan Centre for Neuroscience, Milan, Italy; Department of Laboratory Medicine, ASST “Grande Ospedale Metropolitano” Niguarda, Milan, Italy; Neurology department, ASST “Grande Ospedale Metropolitano” Niguarda, Milano, Italy; Istituto Auxologico Italiano, IRCCS, Bioinformatics and Statistical Genomic Unit, Milano, Italy; University of Bologna, Bologna, Italy, Chair of the “Cognitive and Behavioral Neurology” Study Group of the Italian Neurological Society, Italy; Department of Oncology and Hemato-Oncology, University of Milan, Milan, Italy

**Keywords:** mental fatigue, COVID-19, dysnatraemia, electrolyte imbalance

## Abstract

**Objective:** Coronavirus disease 2019 (COVID-19) may induce short- and long-term cognitive failures after recovery, but the underlying risk factors are still a matter of debate. Identifying patients at the highest risk is now a research priority to prevent persistent symptoms after recovery. In this study, we investigated whether: (i) the odds of experiencing persistent cognitive failures may differ based on the patients’ disease course severity and sex; (ii) the patients’ electrolytic profile at the acute stage may represent a risk factor for persistent cognitive failures.

**Methods:** We analysed data from 204 patients suffering from COVID-19 and hospitalised during the first pandemic wave. According to the 7-point WHO-OS Scale, their disease course was classified as *severe* (if the patient needed ventilation) or *mild* (if they did not). We investigated the presence of persistent cognitive failures using a modified version of the Cognitive Failures Questionnaire, collected after hospital discharge, while electrolyte profiles were collected during hospitalisation. We explored our hypotheses via logistic regression models.

**Results:** Females who suffered from mild COVID-19 were more likely to report mental fatigue than those with severe COVID-19 (*β*= 0.29, *95%CI* [0.06; 0.53], *p*= 0.01). Furthermore, they present a statistically significant risk effect of *Na+* alteration at the acute phase on the odds of presenting persistent mental fatigue (*β*= 0.37, *95%CI* [0.09; 0.64], *p*= 0.01).

**Interpretation:** These findings have important implications for the clinical management of COVID-19 hospitalised patients. Attention should be paid to potential electrolyte imbalances, mainly in females suffering from mild COVID-19.

**Key Points:** *Question:* Do disease severity and sex predict the risk of persistent cognitive failures in COVID-19 hospitalised survivors? Does electrolytic imbalance at the acute phase represent a risk factor for persistent cognitive failures after recovery?

*Findings:* Females who suffered from mild compared to severe COVID-19 had a higher risk of presenting persistent mental fatigue. In this group, dysnatraemia at the acute stage represented a significant risk factor on the odds of showing such a persistent cognitive failure after recovery.

*Meaning:* Sodium levels must be monitored and balanced during hospitalisation of females affected by mild COVID-19 to prevent mental fatigue among the possible short- and long-term effects.

## Introduction

Individuals who recovered from Coronavirus disease 2019 (COVID-19) may experience a plethora of persistent symptoms, i.e., ‘long COVID’ ^1^. According to the UK National Institute for Health and Care Excellence (NICE) guidelines, the term long COVID describes signs and symptoms that continue or develop after acute COVIDLJ19. It includes both ongoing symptomatic COVIDLJ19 (from 4 to 12 weeks) and postLJCOVIDLJ19 syndrome (12 weeks or more). These manifestations may include fatigue, muscle weakness, shortness of breath or cough, as well as joint or chest pain ^2^, implicating multi-organ alterations following the viral infection. A growing body of research has shown that females are more likely to suffer from persistent symptoms after recovery, with a higher likelihood to report persistent fatigue ^3–5^. This evidence sets the state for challenging scientific investigations, as long COVID mainly affects women ^4^, although vulnerability and mortality from acute COVID-19 infection are higher in men ^6^. Studying the different clinical patterns between males and females during the infection could shed new light on this issue.

Recent studies have highlighted sex differences in electrolyte imbalances caused by SARS-CoV-2 at the acute phase ^7^. Moreover, such electrolyte patterns have been associated with COVID-19 disease in hospitalised patients ^8^. In patients with COVID-19, SARS-CoV-2 enters the cells using angiotensin-converting enzyme 2 (ACE2) as a receptor, which is one of the main effectors of the brain Renin-Angiotensin System (RAS). The virus replicates after entry into the cells, and ACE2 gets downregulated. As a result, there is reduced degradation of angiotensin-II, leading to increased aldosterone secretion and subsequent electrolyte imbalance. Such biochemical condition differs between male and female, as sex hormones influence the expression and modulation of the Brain-RAS pathway responses. Intriguingly, evidence from other clinical populations has shown that electrolyte imbalance is associated with cognitive failures. For instance, hypernatremia (i.e., sodium levels higher than normal) is associated with cognitive deficits, especially in the elderly population ^9,10^. Moreover, hyponatremia (i.e., sodium levels lower than normal) may also be associated with adverse cognitive outcomes ^11,12^. Specifically, whereas the consequences of acute hyponatremia may be severe, including permanent disability and death, mild and moderate hyponatremia may cause cognitive failures, such as attentional deficits ^13,14^.

Interestingly, long COVID also includes persistent cognitive difficulties ^15–19^, and the most frequently described involves attentional impairments. A systematic review performed on 57 studies with 250.351 survivors of COVID-19 found difficulties in concentration (23.8% of the patients’ sample) ^16^. Furthermore, these failures can be present in the form of (mental) fatigue, one of the most commonly experienced persistent symptoms after hospitalisation ^20^. Previous studies have suggested a correlation between persistent cognitive failures and disease severity ^18,21,22^. Patients who benefited from invasive ventilation presented with better cognitive status ^22^; however, this topic is a current matter of debate.

Our understanding of the factors underpinning persistent cognitive failures after COVID-19 remains limited. Identifying patients at the highest risk is now a research priority to prevent persistent short and long-term symptoms after recovery. Starting with this clinical and scientific need, the current study aims at exploring whether: (i) the probability of experiencing persistent cognitive failures may differ on the bases of the disease course severity and the patients’ sex; (ii) the patients’ electrolytic profile at the acute stage may represent a risk factor for persistent cognitive failures. To these aims, patients were classified into two groups identifying the disease course severity according to whether patients needed ventilation (Orotracheal Intubation or CPAP ventilation n=73 – *severe* COVID-19) or did not need ventilation during hospitalisation (oxygen therapy or no oxygen therapy at all n=131 – *mild* COVID-19), in accordance with the 7-point WHO-OS Scale. To the first aim, we performed an association analysis of disease course severity on the presence of persistent cognitive failures, testing, through an interaction term, if the patients’ sex at birth would moderate the association. Then, for the second aim, we tested via a logistic regression model whether different electrolyte profiles during the infection may impact the persistent cognitive failures highlighted in the previously described analysis according to the patients’ sex in the two groups of disease course severity. Based on previous findings showing that the disease severity plays a role ^22^, we expected that the odds of presenting cognitive failures would be higher in patients who did not need ventilation therapy in the acute phase (mild COVID-19). Furthermore, as long COVID symptoms are more likely to occur in females ^4^, we expected that patients’ sex might interact with the disease severity in the odds of persistent cognitive difficulties. Lastly, as electrolyte imbalances are one of the recurrent features of COVID-19, and it differs between males and females ^7^, we hypothesised that the odds of cognitive failures could be associated with the electrolyte profile during the hospitalisation, particularly in female patients.

## Materials and method

### Patients

We collected data from 275 consecutive patients suffering from COVID-19 admitted to the *ASST Grande Ospedale Metropolitano Niguarda* in Milan during the first pandemic wave in Italy, from February to April 2020 (T0). The diagnosis was based on at least one positive test result at the reverse transcriptase-polymerase chain reaction (PCR) for SARS-CoV-2. After two consecutive negative oropharyngeal swabs (i.e., recovery), patients were discharged and followed up at the outpatient service of the same hospital from May to July 2020 (T1).

As this study focuses on subjective cognitive failures, patients with previous neurological and psychiatric disorders were excluded (*n*=41). In addition, patients diagnosed with chronic obstructive pulmonary disease (COPD) were excluded to ensure that any cognitive outcomes were not related to previous chronic respiratory illness (*n*=9). Furthermore, those patients who did not complete the questionnaire on cognitive failures (*n*=21) were removed from the sample. The final sample comprised 204 patients with a mean age of 57.1 years (±11.9), 130 (63%) of whom were males. Our sample demographics align with those of the Italian National Institute of Health, which reported that participants who tested positive during this period were on average 58 years old.

Patients were assessed at an average of 49.9 (±16.7) days after recovery (follow-up time). Based on their medical records, patients were classified into two groups according to whether they received Orotracheal Intubation or CPAP ventilation (ventilated patients: n=73) or oxygen therapy or no oxygen therapy at all (non-ventilated patients: n=131), thus creating the covariate severity of COVID-19 course. According to the 7-point WHO-OS Scale, the course of COVID-19 in ventilated patients was classified as severe (*severe COVID-19*), while in non-ventilated patients was classified as mild (*mild COVID-19*). The clinical characteristics of the sample separately in the two groups of disease severity are reported in Table 1. The Ethical Committee Comitato Etico Area 3 Milano approved the study (N92-15032020 and N408-21072020). The study was conducted following the Declaration of Helsinki.

**Table 1.** Demographics and clinical characteristics of the patients’ samples. The interactions between patients’ disease severity and sex for the demographic and clinical variables were not statistically significant.

|  | Mild COVID-19<br>(N=131) |  | Severe COVID-19<br>(N=73) |  | Interaction Disease<br>Severity by Sex |
| --- | --- | --- | --- | --- | --- |
|  | *males* | *females* | *males* | *females* | *p-value* |
| *N* | 78 | 53 | 52 | 21 | 0.096 |
| *Age, mean (SD)* | 58.3 (14.5) | 57.8 (14.3) | 55.0 (11.5) | 62.4 (11.5) | 0.069 |
| *Length of hospital stay, mean (SD)* | 15.5 (9.9) | 15.01 (7.6) | 22.1 (11.1) | 24.7 (13.6) | 0.373 |
| *Follow-up time, mean (SD)* | 53.6 (16.6) | 49.6 (13.6) | 46.8 (17.4) | 45.1 (19.6) | 0.664 |

### Clinical questionnaire on cognitive failures

To explore cognitive failures during the health emergency in April 2020, we decided to adopt a clinical tool that could be at the same time effective and quick to administer. Thus, we used a modified version of the Cognitive Failures Questionnaire ^23^. We adjusted some of the original questions (such as those involving social interaction) to adapt them to the quarantine situation the patients might have experienced once discharged from the hospital. The questionnaire consisted of 19 statements related to possible cognitive failures experienced in everyday life involving several domains, such as attention, memory, gnosis, praxis, orientation in time and space and executive functions (see Table 2 for the complete statements list). Similar questionnaires have also been recently used to test cognitive failures during quarantine/self-isolation for COVID-19 ^24^. In our study, patients were asked to indicate the presence or absence of cognitive failures with a “yes/no” response. They could report more than one symptom. At the follow-up visit, the patients came to the Chronicity Service of the *ASST “Grande Ospedale Metropolitano” Niguarda*, where they underwent a series of assessments throughout the morning. The dedicated healthcare staff administered the cognitive failures questionnaire to all subjects, among several other evaluations.

**Table 2.**
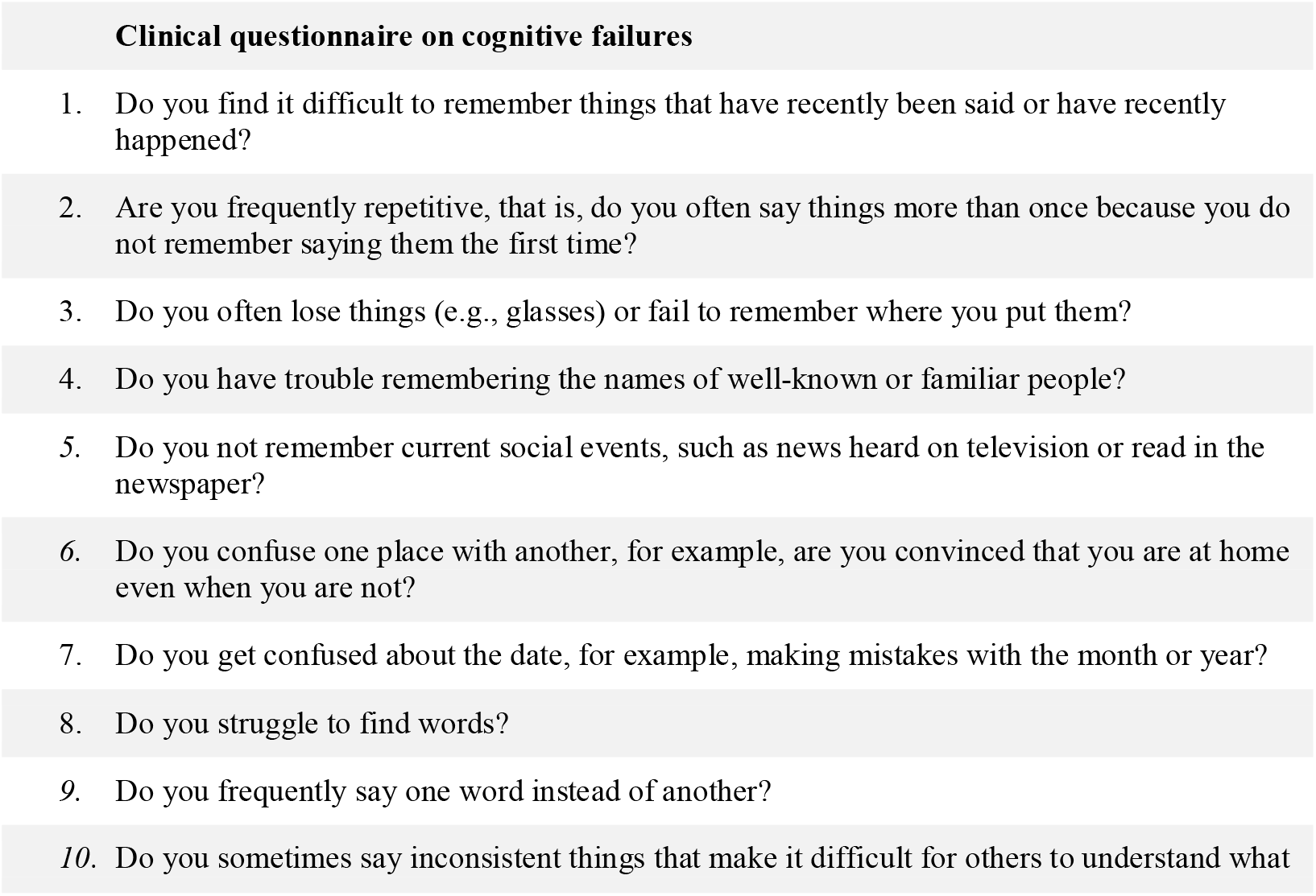
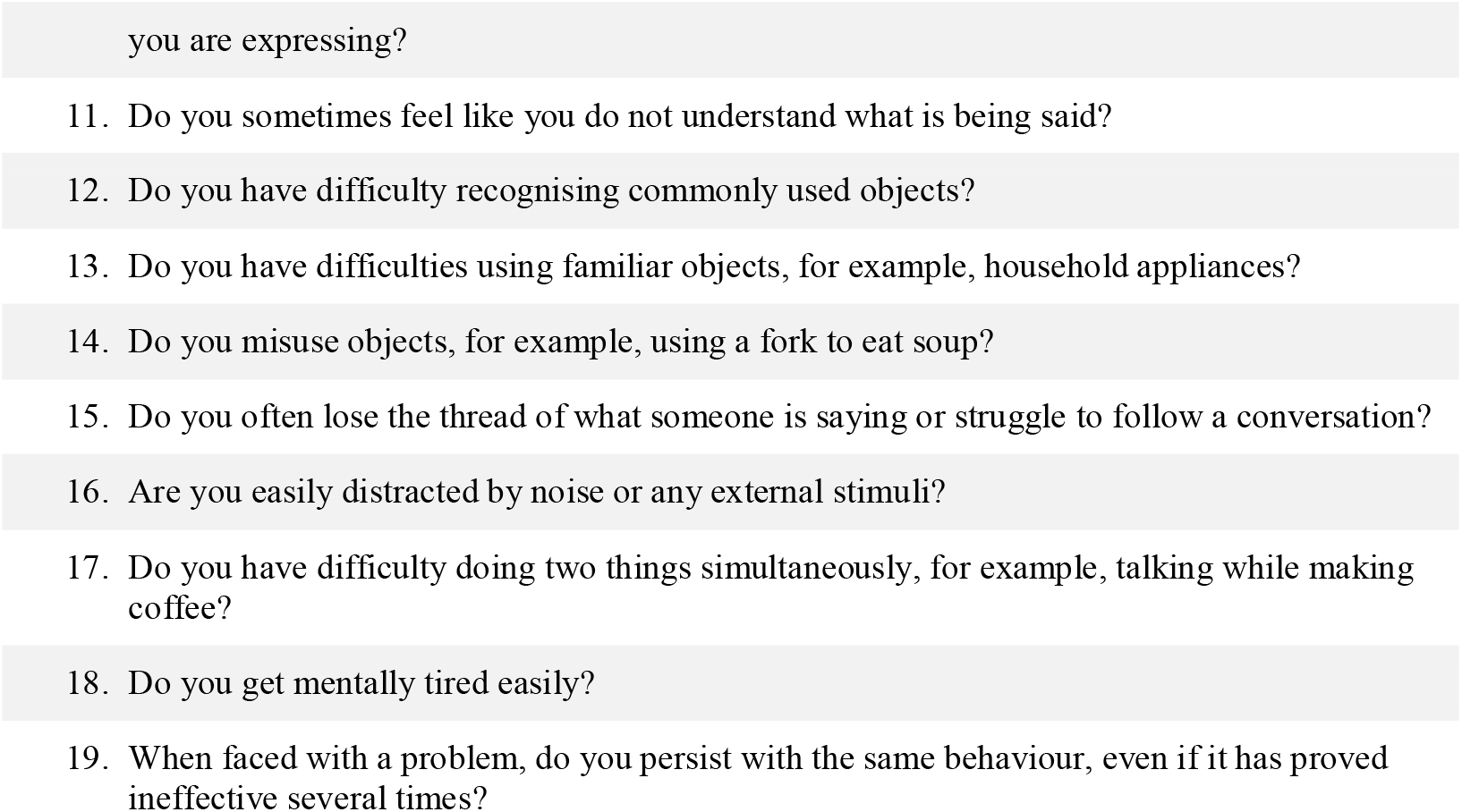
Cognitive failures questionnaire. The table shows the complete list of statements included in the questionnaire. For each statement, patients provided a yes/no response.

### Laboratory test and data sources

We retrospectively extracted the electrolytic profile for each patient from a panel of laboratory test results performed for the clinical routine. The tested Chemical analytes included chloride (*Cl-*), potassium (*K+*), and sodium (*Na*+). All samples were analysed in duplicate, within one hour from blood collection, using the same analyser and the same lot of reagents. Electrolyte parameters were measured by a Roche Cobas 8000 system (ISE modules). Blood samples were processed in a centrifuge at 3000 rpm (revolutions per minute). To obtain laboratory variables, a query was created to extract anonymised data using the patients’ ID (a numeric string) from a SQLLJbased repository in which all analytical results of the tests performed in the laboratory were stored. The fields extracted were sex, date of birth, day of lab tests execution, tests ID, test results and hospital ward.

### Statistical analysis plan

Firstly, we compared the demographic variables reported in Table 1 between the patient groups. We performed a chi-square analysis for categorical variables and t-test or Wilcoxon for the continuous ones. Then, a reliability analysis was carried out on the items included in the cognitive failures questionnaire. As the questionnaire involves dichotomously scored items, we used the Kuder-Richardson formula (KR-20), a widely used method to evaluate internal consistency in cognitive and personality tests.

To explore whether the COVID-19 course severity impacted the odds of experiencing cognitive failure after recovery differently depending on the patient’s sex, a logistic regression model for each item of the questionnaire has been fitted. In each model, disease severity, sex at birth, and the interaction between sex and disease severity group were specified as independent variables. We also decided to include the follow-up visit time in the model as it ranged from 3 to 104 days (mean 49.9 (±16.7) days after recovery). Each item representing a distinct cognitive failure was specified as a dependent variable. If the interaction between course disease severity and sex resulted statistically significant, meaning that the effect of COVID-19 course severity on the investigated endpoint was statistically significantly different in the two sexes, we fitted the logistic regression model as above, stratifying by sex to estimate the effect of COVID-19 course severity in each stratum of the sex variable. If the interaction was not statistically significant, a model as above but with solely the main effects of disease severity group, sex, and follow-up visit was fitted.

Lastly, we tested the hypothesis that the odds of presenting specific persistent cognitive failures after the recovery could be associated with the electrolyte imbalance observed during the hospitalisation. Thus, we fitted logistic regression models considering only the questionnaire items significantly different according to the patients’ sex and disease severity. All the laboratory variables were transformed from continuous into binary variables according to their specific cut-off value: if a variable value was out of the normal range, it was labelled “1”; otherwise, the value was labelled “0”. Importantly, for all the electrolytes, a score equal to 1 means that the variable values are higher or lower than the normal range. For instance, in the case of *Na+*, a score equal to 1 means that patients presented with dysnatraemia (hyponatremia, hypernatremia or both).

Statistical analyses were performed using Jamovi software (version 1.2).

## Results

### Persistent cognitive failures: the impact of the disease course severity and sex

Concerning the socio-demographic and clinical variables, we found no significant difference in terms of interaction between disease course severity and sex for age (*F*_(1,198)_= 3.3, *p*= 0.069), sex (*X*^*2*^_(1)_= 2.8, *p*=0.096), follow-up time (*F*_(1,188)_= 0.2, *p*= 0.664) and hospital length stay (*F*_(1,188)_= 0.8, *p*= 0.373). See Table 1. Concerning the questionnaire, the reliability analysis of the items showed good internal consistency (*KR-20* = 0.851).

Results of the logistic regression analysis aimed at investigating whether the odds of cognitive failures may differ on the bases of the patients’ disease severity and sex showed a statistically significant interaction between COVID-19 course severity and sex (*β*= 0.32, *95%CI* [0.08;0.55]), *p*=0.009) for the mental fatigue (Item 18) only. For this item, we performed a subsequent analysis by stratifying by sex. Results of this latter analysis showed a statistically significant effect of group severity in females (*β*= 0.29, *95%CI* [0.06; 0.53], *p*= 0.01), meaning that females who suffered from a mild course compared to a severe course of COVID-19 have a higher risk to present persistent mental fatigue after recovery. No effect was observed in males (*β*= -0.01, *95%CI* [-0.14; 0.11], *p*= 0.82) (see Table 3).

**Table 3.** Results of the sex-stratified logistic regression model of COVID-19 course severity and cognitive failures, adjusting for follow-up time.

| ITEM 18 | *Sex = Female* |  |  | *Sex=Male* |  |  |
| --- | --- | --- | --- | --- | --- | --- |
| | $\beta$ | 95%CI | p-value | $\beta$ | 95%CI | p-value |
| **Mild COVID-19** | 0.29 | 0.06;0.53 | 0.01 | -0.01 | -0.14; 0.11 | 0.82 |
| **Follow-up time** | 0.004 | -0.003;0.01 | 0.27 | 0.003 | -0.0007; 0.006 | 0.12 |

A logistic regression model, without interaction term, was fitted for all the remaining items, in which no statistically significant interaction was observed. As reported in Supplementary Table 1, no statistically significant effect of COVID-19 group severity was observed for any investigated item.

### Association between persistent mental fatigue and electrolyte profile during hospitalisation

Due to some missing data, we retrospectively analysed laboratory test results from 197 patients (out of 204). The sample was composed by *n*= 125 mild COVID-19 patients (age: *M*=58.08 (± 14.5); sex: 73 males) and *n*= 72 severe COVID-19 patients (age: *M*= 57.21 (± 11.9); sex: 52 males). Results of the logistic regression models performed on Item 18, resulting from the previous analysis, showed a statistically significant risk effect of *Na+* alteration (*β*= 0.37, *95%CI* [0.09; 0.64], *p*= 0.01) on the odds of presenting persistent mental fatigue after recovery in females who suffered from a mild course of COVID-19. No statistically significant effects were observed for the remaining electrolytes in females with mild disease course: *K+* (*β*= 0.01, *95%CI* [-0.26; 0.28], *p*= 0.94) and *Cl-* (*β*= -0.04, *95%CI* [-0.33; 0.25], *p*= 0.77).

## Discussion

In Italy, during the first wave of the COVID-19 outbreak in 2020 (February 21^st^–June 28^th^), there were a total of 240,760 confirmed infections with 34,788 deaths. Depending on the disease severity, many symptoms may persist after recovery, mainly affecting women. Among these symptoms, cognitive failures may also be experienced. Little is known about the risk factors underpinning persistent symptoms after the recovery. This study set out the challenge to explore whether cognitive failures after recovery may depend upon the disease severity, the patient’s sex, and the electrolytic indices during the hospitalisation.

We confirm previous evidence by showing that cognitive failures may persist for about one month after recovery. Our findings indicated that the disease severity specifically impacted the attentional system by showing that the occurrence of persistent mental fatigue was higher in those patients who suffered from a mild course of COVID-19 (i.e., non-ventilated patients). This result is in line with the study by Alemanno and colleagues ^22^, in which the authors found better cognitive status in those patients who had undergone invasive (orotracheal) ventilation compared to patients undergoing non-invasive ventilation or no ventilation at all. The authors reported that 12 out of 22 survivors (54.5%) who underwent orotracheal ventilation one month after hospital discharge showed an impaired total score on the MoCA test, a well-known global cognitive screening test. The same applied to 10 out of the 12 survivors who were treated with non-invasive ventilation (83.3%), 17 out of the 20 survivors who needed oxygen therapy (85%), and 2 out of the 2 survivors who did not need oxygen-based treatment (100%). Furthermore, studies exploring cognitive outcomes in COVID-19 survivors reported mental fatigue as one of the most recurrent symptoms ^25,26^. Here we also showed that patients’ sex plays a role in developing persistent cognitive failures. Indeed, females who recovered from a mild compared to severe course of COVID-19 were more likely to experience persistent mental fatigue. This result aligns with previous evidence highlighting the role of patients’ sex in presenting persistent fatigue after recovery. A study on 377 patients has shown that the 69% of the sample presented long COVID. The female sex was independently associated with persistent symptoms, and fatigue was most commonly reported (39.5% of the sample) ^5^. Furthermore, another study has shown that females under 50 reported worse fatigue, more likely than men of the same age to experience fatigue ^4^.

It has been postulated that the aetiology of persistent cognitive failures could derive from the dual action of the viral infection that has both direct (immunological and neurological damage) and indirect (hypoxic/respiratory states) consequences ^27^. Indeed, two primary routes are used to explain cognition-related deficits in COVID-19 survivors: virus-induced CNS damage (neurotrophic) or non-CNS impairments ^28^. Strikingly, we found that in females who suffered from a mild course of COVID-19, persistent mental fatigue was related to electrolyte imbalance during hospitalisation. The general symptom of fatigue has been previously reported as a consequence of hyponatremia conditions in hospitalised patients ^29^. In the case of COVID-19, a possible explanation for the correlation between electrolyte imbalance and mental fatigue in females could be represented by the different sex-dependent expressions of the brain Renin-Angiotensin System (RAS). In fact, the expression and the modulation of the Brain-RAS pathway response are influenced by sex hormones. Indeed, with estrogenic stimulation, the ACE2/Ang-(1-7)/MasR system and the ACE2/Ang-(1-8)/AT2 system is increased, while testosterone stimulation mediates the activation of the ACE/Ang- (1-8)/AT1R arm of the RAS. As SARS-CoV-2 enters the cells using ACE2 as a receptor, which is one of the main effectors of the brain RAS, sex differences in the electrolyte imbalance during COVID-19 could be present (Figure 1).

**Figure 1.**
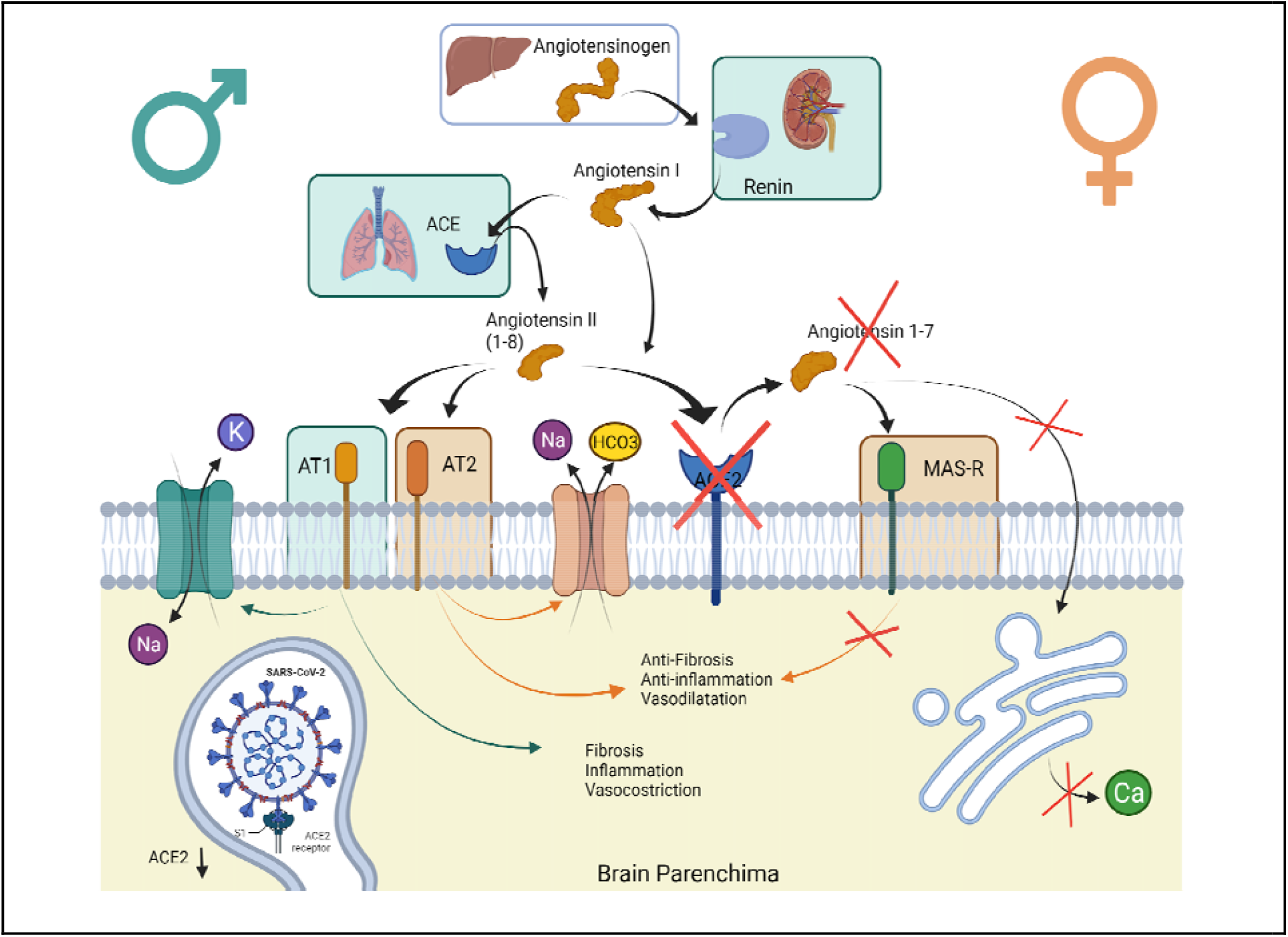
The sex difference in RAS and response of SARS-CoV-2 injury. Several studies revealed that the AT1 receptor protein is down-regulated by oestrogen, while AT2 receptors are up-regulated. On the other hand, testosterone-induced the expression of AT1. AT1 and AT2 receptors have antagonistic action: Sodium cellular intake is mediated by the AT1 receptor through the increase in Na+/K+ ATPase activity, while the AT2 receptor activates phospholipase A2, which contributes to the activation of the Na /HCO3 symporter system (NBC), mediating the Sodium cell excretion.

## Data Availability

All data produced in the present study are available upon reasonable request to the authors

## Conclusions and clinical significance

In summary, this study provided new evidence on the aetiological nature of persistent mental fatigue in COVID-19 survivors who required hospitalisation. In particular, females who suffered from a mild course of COVID-19 presented a higher frequency of reporting this symptom a month after the hospital discharge. If, on the one hand, epidemiological studies reported a lower frequency of hospitalisation for females during the COVID-19 infection, on the other hand, electrolyte imbalance occurs more frequently in this population, possibly causing such a persistent cognitive failure. These results pave the way for a specific electrolyte rebalancing treatment for hospitalised COVID-19 females who do not require ventilation to prevent cognitive failures after recovery. Therefore, adequate laboratory monitoring and subsequent review of appropriate intravenous water balance medications are important management aspects.

## Data and materials availability

The corresponding author will provide the data supporting this study upon reasonable request.

## Credit authorship contribution statement

Conceptualisation, Methodology, Formal analysis, Writing - Review & Editing, Writing - Original Draft: GS, FC, DC, AB, SB, EI; Formal analysis Writing - Original Draft, Visualization, Data Curation: EI, LB, GS; Investigation, & Writing - Review & Editing: FV, GP, AB, SB, GS; Writing - Review & Editing, Conceptualisation: MG, GG, DG, LB, AS, FS, ECA, GB; Supervision, Project administration: GS, GB. All authors have read and agreed to the published version of the manuscript.

