## Supplementary Table 1 for "The association between dysnatraemia during hospitalisation and post COVID-19 mental fatigue"

**Supplementary Table 1**. Results of the logistic regression model for the association between COVID-19 severity group and cognitive failures as investigated by different items. Models were corrected by sex, and follow-up time.

| Cognitive failure | β | SE | *p*-value |
| --- | --- | --- | --- |
| ITEM 1  COVID-19 group severity (mild)  Sex (female)  Follow-up time | 0.06  0.12  0.001 | 0.05  0.05  0.002 | 0.24  0.03  0.49 |
| ITEM 2  COVID-19 group severity (mild)  Sex (female)  Follow-up time | 0.05  0.18  -0.002 | 0.04  0.04  0.001 | 0.22  <0.0001  0.09 |
| ITEM 3  COVID-19 group severity (mild)  Sex (female)  Follow-up time | 0.02  0.13  -0.001 | 0.06  0.06  0.002 | 0.78  0.03  0.57 |
| ITEM 4  COVID-19 group severity (mild)  Sex (female)  Follow-up time | -0.03  0.12  -0.01 | 0.05  0.05  0.0008 | 0.50  0.02  0.57 |
| ITEM 5  COVID-19 group severity (mild)  Sex (female)  Follow-up time | 0.04  0.06  0.0008 | 0.04  0.04  0.001 | 0.35  0.11  0.46 |
| ITEM 6  COVID-19 group severity (mild)  Sex (female)  Follow-up time | 0.01  0.03  -0.0001 | 0.02  0.02  0.0004 | 0.36  0.08  0.79 |
| ITEM 7  COVID-19 group severity (mild)  Sex (female)  Follow-up time | 0.07  0.06  -0.0006 | 0.04  0.03  0.001 | 0.06  0.10  0.57 |
| ITEM 8  COVID-19 group severity (mild)  Sex (female)  Follow-up time | 0.04  0.12  0.003 | 0.05  0.05  0.002 | 0.44  0.04  0.11 |
| ITEM 9  COVID-19 group severity (mild)  Sex (female)  Follow-up time | 0.02  0.06  0.002 | 0.03  0.03  0.001 | 0.55  0.09  0.03 |
| ITEM 10  COVID-19 group severity (mild)  Sex (female)  Follow-up time | 0.03  0.05  0.0006 | 0.02  0.02  0.0007 | 0.18  0.05  0.37 |
| ITEM 11  COVID-19 group severity (mild)  Sex (female)  Follow-up time | 0.04  0.17  0.002 | 0.05  0.05  0.001 | 0.45  0.0004  0.26 |
| ITEM 12  COVID-19 group severity (mild)  Sex (female)  Follow-up time | 0.009  0.01  -0.0003 | 0.01  0.01  0.0003 | 0.43  0.25  0.33 |
| ITEM 13  COVID-19 group severity (mild)  Sex (female)  Follow-up time | 0.002  0.02  0.0003 | 0.02  0.02  0.0006 | 0.92  0.30  0.64 |
| ITEM 14  COVID-19 group severity (mild)  Sex (femal  Follow-up time | 0.009  0.01  -0.0003 | 0.01  0.01  0.0003 | 0.43  0.25  0.32 |
| ITEM 15  COVID-19 group severity (mild)  Sex (female  Follow-up time | 0.03  0.15  0.002 | 0.04  0.04  0.001 | 0.45  0.0008  0.14 |
| ITEM 16  COVID-19 group severity (mild)  Sex (female)  Follow-up time | 0.08  0.17  0.0004 | 0.05  0.05  0.002 | 0.15  0.003  0.82 |
| ITEM 17  COVID-19 group severity (mild)  Sex (female)  Follow-up time | 0.04  0.0009  -0.0009 | 0.03  0.03  0.001 | 0.21  0.98  0.35 |
| ITEM 19  COVID-19 group severity (mild)  Sex (female  Follow-up time | 0.03  0.03  0.0007 | 0.03  0.03  0.0008 | 0.33  0.27  0.38 |
